# A method to estimate the incidence of transfusion reactions for transfusion-treated diseases in Chinese hospitals

**DOI:** 10.1101/2020.02.25.20026096

**Authors:** Feng Lin, Xue Tian, Yonghua Yin, Zhong Liu

## Abstract

Although transfusion reactions are directly related to transfusion components, transfusion components are clinically related to the diseases afflicting patients. As some transfusion reactions can be life-threatening, estimating the incidence of transfusion reactions (which, in this study, are strictly defined as allergic and febrile non-haemolytic transfusion reactions) of certain categories of diseases is helpful for clinicians. According to the reported blood use of specific departments, the numbers of transfusion patients in these departments can be estimated. By the Association rule mining algorithm, the categories of diseases that often correlate with transfusion reactions have been screened for. It is found that the diseases belonging to C00-C97, D00-D48, D50-D89, K00-K93, N00-N99 and O00-O99 (ICD-10) often correlate with transfusion reactions. Platelet transfusion patients whose diseases belong to C00-C97 encounter transfusion reactions with an incidence of about 1%, which is much higher than the average. The incidence of transfusion reactions in patients whose diseases belong to K00-K93 –who undergo plasma transfusions –might be higher than the average, as the lower bound of this incidence is equivalent to the average incidence. Based on this study, it is suggested that attention be paid to patients whose diseases belong to C00-C97 –who undergo platelet transfusions –to prevent them from experiencing allergic transfusion reactions.

## 1 Introduction

Transfusions are a kind of therapeutic procedure performed on hospitalized patients [1–4]. It is well known that errors in blood transfusion practices can lead to serious consequences for recipients in terms of morbidity and mortality [5–8]. The morbidity or mortality caused by transfusions are called adverse transfusion reactions or transfusion reactions (TRs). The incidence of transfusion reactions is about 0.2% [9, 10]. Most transfusion reactions are related to immune reactions [11]. Allergic and febrile nonhaemolytic transfusion reactions (FNHTR) are the most common transfusion reactions, and clinicians most often encounter this type. In this study, allergic and FNHTRs constitute a sufficiently large sample size, so in this paper we only focus on these two kinds of transfusion reactions.

For clinicians, attention must be paid to the diseases that afflict their patients. They make decisions about whether their patients need transfusions according to their patients’conditions. As some mechanisms of TR still remain unknown [12–16], estimating the incidence of transfusion reactions corresponding to certain of diseases is better than judging the incidence of transfusion reactions by average incidence. This is helpful for clinicians in preventing transfusion patients from experiencing transfusion reactions.

Though the numbers of transfusion patients in certain disease categories are supplied, they can be roughly estimated by the blood use of various departments. Information on transfusion reactions among patients (including the departments in which they were in, their categories of TRs and the diseases afflicting them) has already been collected in a database. This means that an interval or at least a lower bound for such incidence can be estimated. To screen specific categories of diseases that often correlate with transfusion reactions, Association rule mining algorithm [17–23] is used. According to the numbers of samples that belong to the specific categories of diseases screened by the Association mining rule algorithm, lower bounds or intervals of the incidence of transfusion reactions for specific categories of diseases can be estimated.

## 2 Materials and methods

### 2.1 Ethical approval

All patients involve in the study had given written informed consent by participating hospitals. The current project was approved by the review board of the Research Ethics Committee of Institute of Blood Transfusion, Chinese Academy of Medical Sciences and Peking Union Medical College on September 18, 2016 (registration number 2016017).

### 2.2 Materials

Samples of transfusion reaction patients are supplied by the Key Laboratory of Adverse Transfusion Reactions (the Key Laboratory). The Key Laboratory collects data on clinical cases of transfusion reactions. A total of 79 hospitals have joined the Key Laboratory. Apart from Tibet, the samples supplied by these hospitals are representative of all transfusion reaction patients in the Chinese mainland.

From May 2018 to June 2019, 1255 cases were collected. In this study, the sample size for allergy and FNHTR was large enough for statistical research. Among the 1255 cases, only 839 cases appeared to be allergy or FNHTR. According to the clinical diagnoses, there were 754 confirmed cases. Table 1 shows the number of transfusion patients and the total quantity of transfusion components.

**Table 1:** The total quantity of transfusion patient and blood components

|  | Number | Transfusions | Red<br>blood cells | Plasma | Concentrated<br>platelets | Apheresis<br>platelets | Cryoprecipitation |
| --- | --- | --- | --- | --- | --- | --- | --- |
| Total | 681077 | 1188380 | 1264293.85 | 136100947.38 | 10315.5 | 240190.1 | 1545916.2 |

The data in Table 1 only supply the number of transfusion patients, whereas the numbers of patients who belong to certain categories of diseases that required transfusions have not been supplied. Estimating the incidence of transfusion reactions requires two kinds of information: the number of transfusion patients who belonged to a specific category of diseases and how many of them had transfusion reactions. The former has not been supplied, so it must be estimated. The latter, however, has been supplied by the Key Laboratory. The “Chinese Clinical Blood Use Research Report (2016)”is helpful for making this estimate. This report was completed in 2017, and it reported the proportion of blood use in the departments shown in Table 2.

**Table 2:** Proportion of blood use in each department

| Department | Red blood cells | Plasma | Platelets | Cryoprecipitation |
| --- | --- | --- | --- | --- |
| Internal medicine | 24% | 27% | 48% | 15% |
| Surgery | 39% | 42% | 17% | 23% |
| Gynaecology and obstetrics | 6% | 3% | 4% | 3% |
| ICU | 9% | 15% | 6% | 15% |
| Paediatrics | 3% | 3% | 6% | 34% |
| Others | 19% | 10% | 19% | 8% |

### 2.3 Estimation of the number of transfusion patients

Following Table 2, we first estimated the average blood dosage in each department. With the average dosage, the number of transfusion patients in each department could be estimated. According to the transfusion dosage records supplied by the hospitals, the estimated average transfusion dosages are shown in Table 3.

**Table 3:**
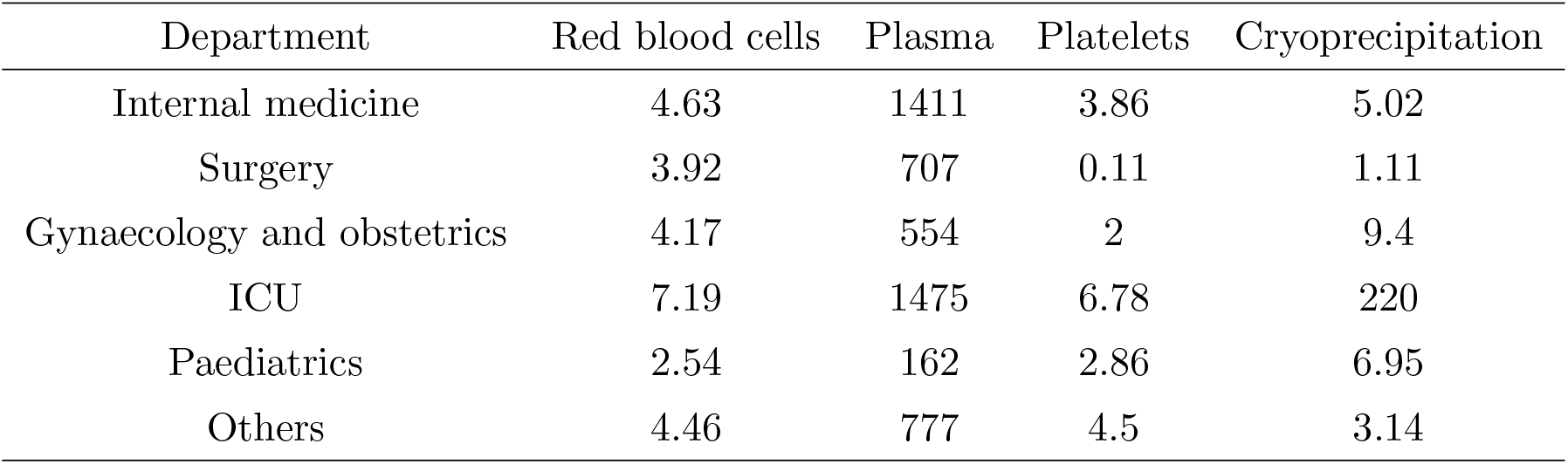
Average dosage estimated in each of the department

According to Table 3 and tables 1-2, the numbers of transfusion patients in each department can be estimated, as shown in Table 4.

**Table 4:** Estimated number of transfusion patients in each of the department

| Department | Red blood cells | Plasma | Platelets | Cryoprecipitation |
| --- | --- | --- | --- | --- |
| Internal medicine | 65535 | 26043 | 31150 | 46192 |
| Surgery | 125784 | 80852 | 387145 | 320324 |
| Gynaecology and obstetrics | 18191 | 7370 | 5010 | 4933 |
| ICU | 15825 | 13840 | 2216 | 1054 |
| Paediatrics | 14932 | 25203 | 5255 | 75627 |
| Others | 53860 | 17516 | 10576 | 39386 |

Table 4 supplies the base information on the numbers of transfusion patients. The total numbers of transfusion patients shown in Table 4 are larger than the total numbers supplied by Table 1. The reason for this is that some patients received transfusions of more than one kind of component, and some transfusions were stopped when acute TRs occurred. These situations led to errors regarding average dosage estimates. The estimated numbers were larger than the numbers of transfusions in Table 1. By calculation, the actual number of transfusions was 0.85 times the estimated number of transfusions. Though we are not able to know the exact numbers of transfusion patients who were being treated for certain diseases, the numbers of transfusion patients in each of the departments already give intervals on the numbers of transfusion patients who were suffering certain categories of diseases.

### 2.4 Estimation of the incidence of transfusion reaction

By intuition, some categories of diseases result in transfusion reactions more frequently. The transfusion component used is associated with the patient’s condition, and the patient’s condition is related to the disease for which the patient is being treated. The causality is as such: the disease that the patient is being treated for determines the transfusion component that the clinician chooses to use, and the transfusion component and the patient’s condition lead to certain transfusion reactions. The causes that lead to transfusion reactions form a chain of causality. For each TR patient, a chain of causality can be determined.

Transfusion patients are divided into groups according to the department classifications in tables 2-4. The causes of each of the samples are divided into four parts: the disease that the patient was being treated for, the reason for transfusion, the immune history of the patient and the transfusion component that the patient received. Information on all samples was supplied by the Key Laboratory. A schematic graph of the relation of causes is shown in Figure 1

**Figure 1:**
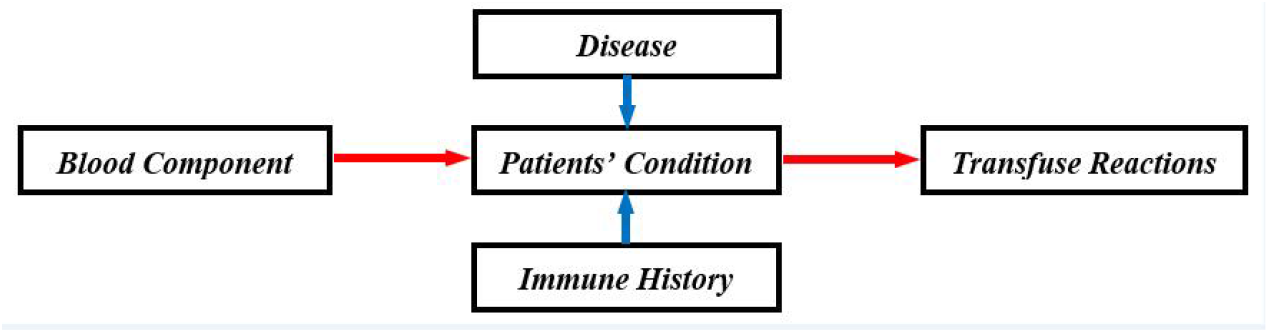
The causality network of transfusion reaction

The support and confidence of the combination of these causes can be determined by the Apriori algorithm. The larger the support is, the stronger the association with the disease and transfusion reaction. Collecting frequently encountered combinations of causes, it is possible to estimate the incidence of transfusion reactions for a certain category of diseases.

The minimal support in the Apriori algorithm is set to 10^*−*4^. This means that as long as a chain of causality is found, this chain of causality is considered to be associated with transfusion reactions. But in practice we choose a minimum support as follows: if the support of the chain of causality is lower than 0.05 in the combination of these four causes, or if it is lower than 0.04 in a combination of four causes plus a result, this causality chain is not taken into consideration. The reason is that the sample sizes of transfusion reactions in each of the departments are not as large as the sample sizes of customer purchasing behavior. Though a low support of a certain kind of combination might indicate a causality chain, it is difficult to conclude that such a combination is of statistical significance.

## 3 Results

The estimated incidence of transfusion reactions in each department is shown in tables 5-10. The classification of diseases accords with the International Classification of Diseases, 10th revision (ICD-10). By Association rule mining algorithm, seven kinds of diseases that would frequently result in transfusion reactions were found. According to reference [24–27] (figure 3 in [28]), the incidence for cancer in China is 0.2%. In the department of internal medicine, according to the records supplied by the Key Laboratory, of all 408 patients that had transfusion reactions (allergy or FNHTR), 137 were cancer patients. It is reasonable to estimate that in the department of internal medicine, 20% of all patients are cancer patients. And according to the data supplied by the Key Laboratory, in the department of internal medicine, nearly 1*/*3(33*outof* 109) of the patients whose diseases belonged to D50-D89 also had cancer. That is why an upper bound for the incidence of transfusion reactions with diseases belonging to C00-C99 and D50-D89 can be estimated in the department of internal medicine.

**Table 5:** The incidence of transfusion reaction in the internal department

| ICD | Lower bound<br>of incidence | Upper bound<br>of incidence | Component | Category<br>of reaction |
| --- | --- | --- | --- | --- |
| C00-C97 | 0.0020 | 0.0143 | Platelets | Allergy |
| D50-D89 | 0.00096 | 0.0026 | Platelets | Allergy |
| K00-K93 | 0.0019 | - | Plasma | Allergy |

**Table 6:** The incidence of transfusion reaction in the surgery department

| ICD | Lower bound<br>of incidence | Upper bound<br>of incidence | Component | Category<br>of reaction |
| --- | --- | --- | --- | --- |
| K00-K93 | 0.0001 | - | Plasma | Allergy |
| S00-T98 | $9.89 \times 10^{-5}$ | - | Plasma | Allergy |

Remarks: In Table 5, myelosuppression after chemotherapy can result in thrombocytopenia, so the range of ATR patients belonging to C00-C97 is estimated as (64, 89). Similarly, the range for ATR patients belonging to D50-D89 is (30, 49) (according to Table 11). The total numbers of transfusion patients who received transfusions of a certain component are shown in Table 4. It has been assumed that 20% of the patients in the department of internal medicine belonged to C00-C93. And since 1/3 of the patients whose diseases belonged to D50-D89 also had cancer, it could be assumed that 0.3 *×* 20% = 6% of the patients in the department of internal medicine belonged to D50-D89. In Table 7,bleeding would result in anemia (according to Table 13), so the range for ATR patients belonging to O00-D99 is (12, 15), but the patients who needed plasma transfusions in the gynecology and obstetrics department did not all belong to O00-O99. So, the upper bound for this incidence could not be estimated. In Table 9, the lower bound for the number of cancer patients is estimated as 53. Although, according to Table 15, bleeding is related to thrombocytopenia, according to the raw data supplied by the Key Laboratory, only one sample showed both bleeding and thrombocytopenia. In Table 10, it is also assumed that 20% of the patients in this department belonged to C00-C93.

**Table 7:** The incidence of transfusion reaction in the gynaecology and obstetrics department

| ICD | Lower bound<br>of incidence | Upper bound<br>of incidence | Component | Category<br>of reaction |
| --- | --- | --- | --- | --- |
| D00-D48 | 0.0001 | - | Red blood cells | FNHTR |
| N00-N99 | 0.0001 | - | Red blood cells | FNHTR |
| O00-O99 | 0.0001 | - | Red blood cells | FNHTR |
| D50-D89 | 0.0001 | - | Red blood cells | Allergy |
| O00-O99 | 0.0003 | - | Red blood cells | Allergy |
| O00-O99 | 0.0016 | - | Plasma | Allergy |

**Table 8:** The incidence of transfusion reaction in the ICU department

| ICD | Lower bound<br>of incidence | Upper bound<br>of incidence | Component | Category<br>of reaction |
| --- | --- | --- | --- | --- |
| K00-K93 | 0.0002 | - | Red blood cells | Allergy |
| K00-K93 | 0.0004 | - | Plasma | Allergy |

**Table 9:** The incidence of transfusion reaction in the paediatrics department

| ICD | Lower bound<br>of incidence | Upper bound<br>of incidence | Component | Category<br>of reaction |
| --- | --- | --- | --- | --- |
| C00-C97 | 0.01 | - | Platelets | Allergy |
| D50-D89 | 0.002 | - | Platelets | Allergy |

**Table 10:**
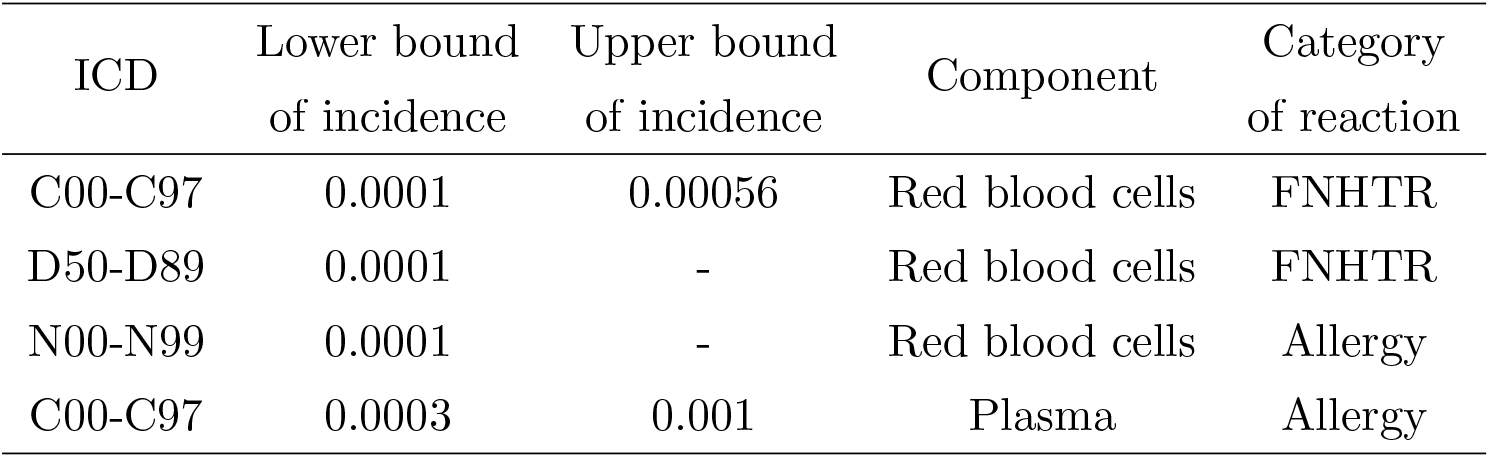
The incidence of transfusion reaction in the other department

**Table 11:**
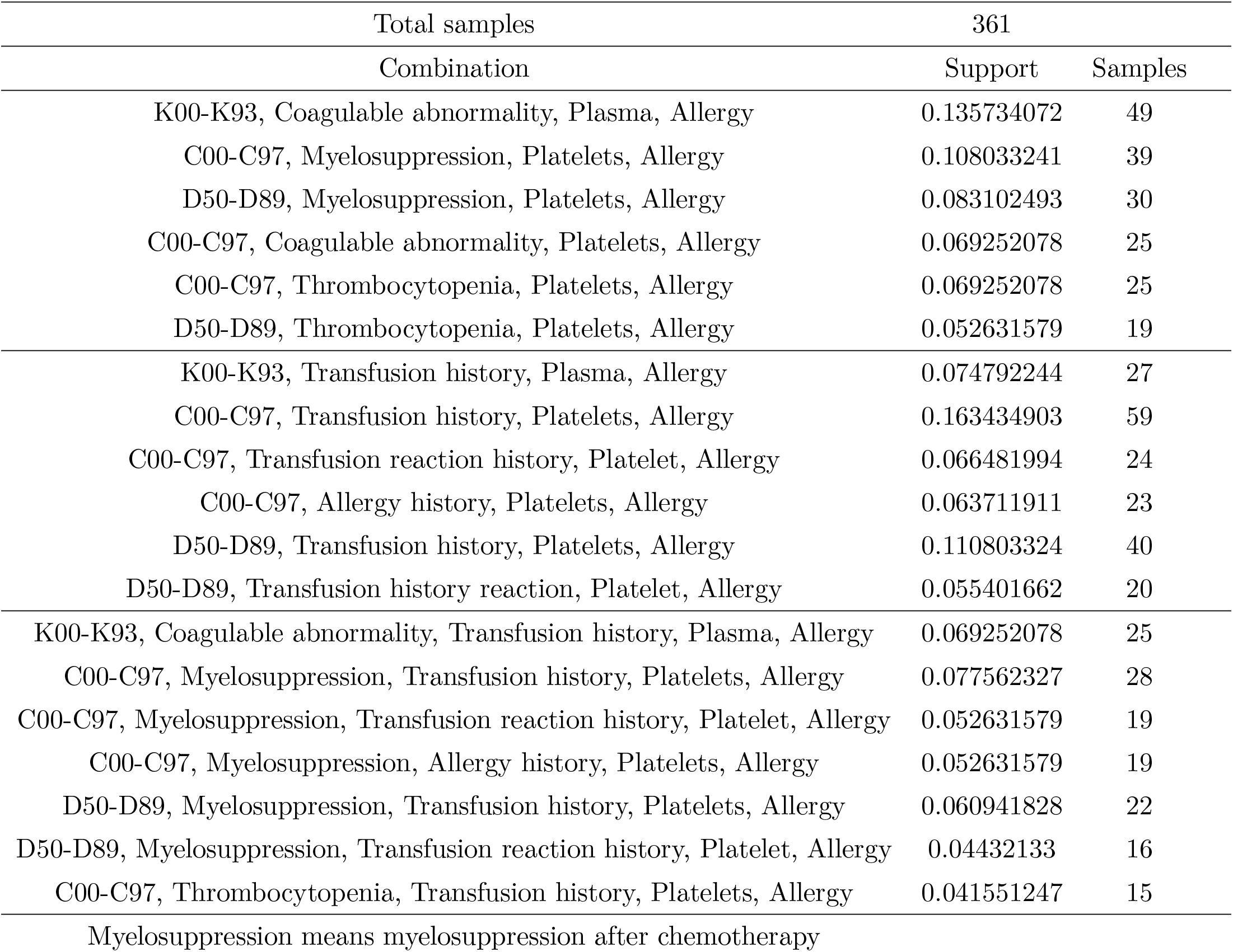
The support of the combination of the causes in the internal medicine department

| Total samples | 361 |  |
| --- | --- | --- |
| Combination | Support | Samples |
| K00-K93, Coagulable abnormality, Plasma, Allergy | 0.135734072 | 49 |
| C00-C97, Myelosuppression, Platelets, Allergy | 0.108033241 | 39 |
| D50-D89, Myelosuppression, Platelets, Allergy | 0.083102493 | 30 |
| C00-C97, Coagulable abnormality, Platelets, Allergy | 0.069252078 | 25 |
| C00-C97, Thrombocytopenia, Platelets, Allergy | 0.069252078 | 25 |
| D50-D89, Thrombocytopenia, Platelets, Allergy | 0.052631579 | 19 |
| K00-K93, Transfusion history, Plasma, Allergy | 0.074792244 | 27 |
| C00-C97, Transfusion history, Platelets, Allergy | 0.163434903 | 59 |
| C00-C97, Transfusion reaction history, Platelet, Allergy | 0.066481994 | 24 |
| C00-C97, Allergy history, Platelets, Allergy | 0.063711911 | 23 |
| D50-D89, Transfusion history, Platelets, Allergy | 0.110803324 | 40 |
| D50-D89, Transfusion history reaction, Platelet, Allergy | 0.055401662 | 20 |
| K00-K93, Coagulable abnormality, Transfusion history, Plasma, Allergy | 0.069252078 | 25 |
| C00-C97, Myelosuppression, Transfusion history, Platelets, Allergy | 0.077562327 | 28 |
| C00-C97, Myelosuppression, Transfusion reaction history, Platelet, Allergy | 0.052631579 | 19 |
| C00-C97, Myelosuppression, Allergy history, Platelets, Allergy | 0.052631579 | 19 |
| D50-D89, Myelosuppression, Transfusion history, Platelets, Allergy | 0.060941828 | 22 |
| D50-D89, Myelosuppression, Transfusion reaction history, Platelet, Allergy | 0.04432133 | 16 |
| C00-C97, Thrombocytopenia, Transfusion history, Platelets, Allergy | 0.041551247 | 15 |

Myelosuppression means myelosuppression after chemotherapy

**Table 12:** The support of the combination of the causes in the surgery department

| Total samples | 126 |  |
| --- | --- | --- |
| Combination | Support | Samples |
| K00-K93, Coagulable abnormality, Plasma, Allergy | 0.079365079 | 10 |
| S00-T98, Coagulable abnormality, Plasma, Allergy | 0.063492063 | 8 |

**Table 13:** The support of the combination of the causes in the gynaecology and obstetrics department

| Total samples | 40 |  |
| --- | --- | --- |
| Combination | Support | Samples |
| D00-D48, Anemia, Red blood cells, Allergy | 0.05 | 2 |
| O00-O99, Anemia, Red blood cells, Allergy | 0.15 | 6 |
| O00-O99, Coagulable abnormality, Plasma, Allergy | 0.175 | 7 |
| O00-O99, Anemia, Plasma, Allergy | 0.05 | 2 |
| O00-O99, Perioperative blood transfusion, Plasma, Allergy | 0.075 | 3 |
| O00-O99, Bleeding, Plasma, Allergy | 0.075 | 3 |
| D50-D89, Anemia, Red blood cells, FNHTR | 0.05 | 2 |
| N00-N99, Anemia, Red blood cells, FNHTR | 0.05 | 2 |
| O00-O99, Anemia, Red blood cells, FNHTR | 0.05 | 2 |
| D00-D48, Transfusion history, Red blood cells, Allergy | 0.05 | 2 |
| D00-D48, Transfusion reaction history, Red blood cells, Allergy | 0.05 | 2 |
| O00-O99, Pregnancy history, Red blood cells, Allergy | 0.075 | 3 |
| O00-O99, Transfusion history, Plasma, Allergy | 0.125 | 5 |
| O00-O99, Pregnancy history, Plasma, Allergy | 0.325 | 13 |
| O00-O99, Pregnancy history, Red blood cells, FNHTR | 0.075 | 3 |
| O00-O99, Pregnancy history, Plasma, FNHTR | 0.05 | 2 |
| D00-D48, Anemia, Transfusion history, Red blood cells, Allergy | 0.05 | 2 |
| D00-D48, Anemia, Transfusion history reaction, Red blood cells, Allergy | 0.05 | 2 |
| O00-O99, Anemia, Pregnancy history, Red blood cells, Allergy | 0.075 | 4 |
| O00-O99, Coagulable abnormality, Transfusion history, Plasma, Allergy | 0.1 | 4 |
| O00-O99, Coagulable abnormality, Pregnancy history, Plasma, Allergy | 0.15 | 6 |
| O00-O99, Perioperative blood transfusion, Pregnancy history, Plasma, Allergy | 0.075 | 3 |
| O00-O99, Bleeding, Pregnancy history, Plasma, Allergy | 0.05 | 2 |

**Table 14:** The support of the combination of the causes in the ICU department

| Total samples | 55 |  |
| --- | --- | --- |
| Combination | Support | Samples |
| K00-K93, Coagulable abnormality, Red blood cells, Allergy | 0.054545455 | 3 |
| K00-K93, Coagulable abnormality, Plasma, Allergy | 0.090909091 | 5 |
| K00-K93, Transfusion history, Red blood cells, Allergy | 0.054545455 | 3 |
| K00-K93, Transfusion history, Plasma, Allergy | 0.054545455 | 3 |
| K00-K93, Pregnancy history, Plasma, Allergy | 0.054545455 | 3 |
| K00-K93, Coagulable abnormality, Transfusion history, Plasma, Allergy | 0.054545455 | 3 |

**Table 15:** The support of the combination of the causes in the paediatrics department

| Total samples | 88 |  |
| --- | --- | --- |
| Combination | Support | Samples |
| C00-C97, Coagulable abnormality, Platelets, Allergy | 0.238636364 | 21 |
| D50-D89, Coagulable abnormality, Platelets, Allergy | 0.102272727 | 9 |
| C00-C97, Thrombocytopenia, Platelets, Allergy | 0.170454545 | 15 |
| C00-C97, Bleeding, Platelet, Allergy | 0.204545455 | 18 |
| C00-C97, Transfusion history, Platelets, Allergy | 0.465909091 | 41 |
| C00-C97, Transfusion reaction history, Platelets, Allergy | 0.136363636 | 12 |
| C00-C97, Allergy history, Platelets, Allergy | 0.102272727 | 9 |
| D50-D89, Transfusion history, Platelets, Allergy | 0.159090909 | 14 |
| C00-C97, Coagulable abnormality, Transfusion history, Platelets, Allergy | 0.227272727 | 20 |
| C00-C97, Coagulable abnormality, Transfusion reaction history, Platelets, Allergy | 0.090909091 | 8 |
| C00-C97, Coagulable abnormality, Allergy history, Platelets, Allergy | 0.079545455 | 7 |
| D50-D89, Coagulable abnormality, Transfusion history, Platelets, Allergy | 0.090909091 | 8 |
| C00-C97, Thrombocytopenia, Transfusion history, Platelets, Allergy | 0.079545455 | 7 |
| C00-C97, Bleeding, Transfusion history, Platelets, Allergy | 0.147727273 | 13 |
| D50-D89, Bleeding, Transfusion history, Platelets, Allergy | 0.045454545 | 4 |

**Table 16:** The support of the combination of the causes in the other department

| Total samples | 84 |  |
| --- | --- | --- |
| Combination | Support | Samples |
| N00-N99, Anemia, Red blood cells, Allergy | 0.095238095 | 8 |
| C00-C97, Coagulable abnormality, Plasma, Allergy | 0.05952381 | 5 |
| C00-C97, Anemia, Red blood cells, FNHTR | 0.071428571 | 6 |
| D50-D89, Anemia, Red blood cells, FNHTR | 0.071428571 | 6 |
| N00-N99, Pregnancy history, Red blood cells, Allergy | 0.095238095 | 8 |
| C00-C97, Pregnancy history, Plasma, Allergy | 0.05952381 | 5 |
| C00-C97, Transfusion history, Platelets, Allergy | 0.095238095 | 8 |
| C00-C97, Pregnancy history, Platelets, Allergy | 0.095238095 | 8 |
| C00-C97, Transfusion history, Red blood cells, FNHTR | 0.05952381 | 5 |
| N00-N99, Anemia, Transfusion history, Red blood cells, Allergy | 0.047619048 | 4 |
| N00-N99, Anemia, Pregnancy history, Red blood cells, Allergy | 0.083333333 | 7 |
| C00-C97, Coagulable abnormality, Pregnancy history, Plasma, Allergy | 0.05952381 | 5 |
| C00-C97, Anemia, Transfusion history, Red blood cells, FNHTR | 0.05952381 | 5 |
| D50-D89, Anemia, Transfusion history, Red blood cells, FNHTR | 0.047619048 | 4 |
| D50-D89, Anemia, Pregnancy history, Red blood cells, FNHTR | 0.047619048 | 4 |

Detailed information on the supports and combinations of causes are shown in tables 11-16, which also show the supports of combinations of causes in each department. These tables supply support information for tables 5-10. According to tables 11-16, apart from the gynecology and obstetrics department, the diseases that belong to C00-C97, K00-K93 and D50-D89 result in transfusion reactions frequently. The transfusion components that most probably cause allergies are platelets and plasma. FNHTR caused by the red blood cell component mainly occurs in the gynecology and obstetrics department.

The guidelines on transfusion reactions [8, 29–31] mention that immune history, especially transfusion history, is related to transfusion reactions, and the causality chains found in tables 11-16 are consistent with that view. Patients whose diseases belonged to the classifications C00-C97, D50-D89 or K00-K93 often received transfusions. That these diseases result in repeated transfusions may be a cause of these transfusion reactions.

## 4 Conclusion and Discussion

As has already been discussed in Section 3, the diseases that frequently correlated with transfusion reactions most probably belonged to the following classifications: C00-C97, K00-K93 and D50-D89. According to tables 5-10, the incidence of transfusion reactions is high in the group of cancer patients, especially when platelets are transfused. In the pediatrics department, transfusion reactions among cancer patients were nearly ten times the average incidence. Patients whose diseases belonged to the classifications C00-C97, D50-D89 or K00-K93 may have received repeated transfusions. That these diseases resulted in repeated transfusions may have been a cause of these transfusion reactions. This result is consistent with the previous study.

Unlike a study seeking to discover information about mechanisms of action via experimental design, the information from this study was extracted from a database. This information shows that attention should be paid to cancer patients who need platelet transfusions. So far, the sample sizes for transfusion reactions are not large. Provided sample sizes are large enough, information on transfusion reactions, such as transfusion-associated circulatory overload and transfusion-related acute lung injury, which might be related to certain kinds of diseases, can be extracted in a similar fashion. This study only demonstrated a method to estimate the incidence of transfusion reactions. If information on the numbers of transfusion patients who were treated for certain kinds of diseases becomes known, the estimates can be made more accurate.

## Data Availability

The data in the manuscript are supplied by the Key laboratory of transfusion adverse reactions Chinese academy of medical science and Peking union medical college

